# Willing but Unable: Physicians’ Referral Knowledge as Barriers to Abortion Care

**DOI:** 10.1101/2021.11.24.21266817

**Authors:** Elizabeth M. Anderson, Sarah K Cowan, Jenny A. Higgins, Nicholas B. Schmuhl, Cynthie K Wautlet

**Affiliations:** Indiana University, Sociology; New York University, Sociology; University of Wisconsin School of Medicine and Public Health, Department of Obstetrics and Gynecology; University of Wisconsin School of Medicine and Public Health, Population Health Institute; University of Colorado Anschutz Medical Campus, Department of Obstetrics and Gynecology

## Abstract

Abortion care is a crucial part of reproductive healthcare. Nevertheless, its availability is constrained by numerous forces, including care referrals within the larger healthcare system. Using a unique study of physician faculty across multiple specialties, we examine the factors associated with doctors’ ability to refer patients for abortion care among those who were willing to consult in the care of a patient seeking an abortion (N=674). Even though they were willing to refer a patient for an abortion, half (53%) of the physicians did not know how and whom to make those referrals, though they care for patients who may need them. Those with the least referral knowledge had not been taught abortion care during their medical training and were in earlier stages of their career than those who had more knowledge. This research exposes another obstacle for those seeking an abortion, a barrier that would be overcome with a clear and robust referral system within and across medical specialties.

## 1. INTRODUCTION

Over 600,000 abortions occur in the United States annually, making abortion a central part of reproductive care in the United States (Kortsmit et al., 2020). By the age of 45, almost a quarter (24%) of women have had an abortion (Jones & Jerman, 2017)^1^. Despite the overall safety of abortion care (Henderson et al., 2005; Raymond & Grimes, 2012), patients and providers face elaborate barriers to receiving and providing care (Cohen & Joffe, 2021; Fuentes & Jerman, 2019; Kavanaugh et al., 2019). Here, we illuminate an under-examined barrier: physician’s ability to refer patients who need abotion services. A referral facilitates abortion access and the inability to provide one, a barrier.

Scholarship on obstacles to abortion care often focuses on policy-related hurdles. In this paper, we examine a different set of structural impediments to abortion by asking: what factors are associated with physician’s ability to refer patients for abortion care? To answer this question, we use a study of physician faculty at the University of Wisconsin School of Medicine and Public Health, the largest medical school in a state with restrictive abortion laws and a “hostile” designation in terms of abortion access (Guttmacher Institute, 2021a). Physicians, like the general public, have varying views on abortion that may or may not impact their medical practice. We focus here only on those physicians who are willing to consult on abortion care to identify systemic, rather than attitudinal, barriers.

### 1.1 Abortion Access in the United States

Abortion is a common and safe medical procedure (Henderson et al., 2005; Jones et al., 2019; Raymond & Grimes, 2012). However, people seeking an abortion in the United States often have to navigate several obstacles before receiving care (Cohen & Joffe, 2021). About two in five women live in counties without a facility that provides abortion, and, on average, women travel 34 miles one way to the facility where they receive an abortion (Fuentes & Jerman, 2019; Jones et al., 2019). Legal regulations are among the most well-known barriers abortion. In states hostile to abortion rights, legal regulations make it more difficult for abortion clinics to operate through unwieldy and medically unnecessary requirements (Guttmacher Institute, 2021b).

Understanding the full landscape of barriers that people seeking an abortion face is crucial, as these barriers exacerbate existing disparities in access to abortion (Dehlendorf & Weitz, 2011). The majority of people who have abortions are non-white and lower income (Jerman et al., 2016), therefore increasing restrictions disproportionately burdens people with fewer resources. Although information about abortion is available online, misinformation is prolific (Patev & Hood, 2021). As a stigmatized event (Cowan, 2017; Kumar et al., 2009; Norris et al., 2011), some people may prefer to seek information about abortions from healthcare providers, rather than family or friends. Consequently, medical providers can be a crucial source of information for those who need an abortion (Kavanaugh et al., 2019).

### 1.2 Abortion in Mainstream Medicine

However, the isolation of abortion from mainstream medicine inhibits providers’ knowledge about abortion. While a primary care service, abortion does not a constitute a part of standard primary healthcare. Rather, abortion services are largely siloed in stand-alone clinics. Around 60% of abortions in 2017 were performed in clinics specializing in abortion and 35% of abortions were performed in non-specialized clinics rather than within physician practices or hospitals (Freedman, 2010; Jones et al., 2019). The marginalization of abortion into clinics results in a professional isolation between abortion providers and those who practice in mainstream medicine (Freedman, 2010).

The marginalization of abortion in medicine is further reflected in its presence (or lack thereof) in medical training, in which abortion-related material is often lacking. Lectures or discussions about abortion are not regularly included in medical student’s pre-clinical education and less than half of programs surveyed reported offering clinical experience on abortion during third year rotations (Espey et al., 2005). Moreover, despite family medicine practitioners’ involvement in reproductive healthcare, only a small proportion of all family medicine residency programs offer abortion training (Greenberg & Nothnagle, 2018; Steinauer et al., 1997)

Even in obstetrics and gynecology (ob-gyn) residency programs, training on abortion care is not universal. The Accreditation Council for Graduate Medical Education (ACGME) has required “opt-out” abortion training for all ob-gyn residents since 1996, meaning that abortion training should be built in as a standard part of an ob-gyn residency program unless the resident elects not to participate (Steinauer et al., 2018). Despite this long-standing requirement, only about two-thirds of the ob-gyn residency programs surveyed in 2014 reported having an opt-out abortion training program with dedicated time for training (Steinauer et al., 2018). Training on abortion during medical residency is crucial for increasing the availability of abortion care as previous research has indicated that completing an ob-gyn residency program with abortion training is associated with a higher likelihood of the physician providing abortions post-residency (Steinauer et al., 2003).

Nevertheless, physicians who are trained and willing to provide abortion care face several institutional and professional barriers toward doing so (Freedman, 2010; Freedman et al., 2010; Stulberg et al., 2016). While some physicians report explicit prohibitions from providing abortions either as a result of employer or institutional rules, others report being dissuaded from providing abortions to prevent conflict with colleagues (Bennett et al., 2020; Freedman et al., 2010).

### 1.3 Abortion Referrals

The structural position of abortion within American medicine as a service that is both challenging to access and requires specialized expertise elevates the importance of referrals for abortion care (Cohen & Joffe, 2021; Kavanaugh et al., 2019). The American College of Obstetrics and Gynecology (ACOG) asserts that all patients should receive referrals, even if the provider is personally opposed (ACOG 2021). In practice, previous studies of ob-gyn and primary care providers indicate that the provision of referrals is not universal (Daniel et al., 2020; Desai et al., 2018; Dodge et al., 2018; Holt et al., 2017; Homaifar et al., 2017; Stulberg et al., 2016). One study of primary care physicians found that, while about two-thirds of physicians regularly referred patients for abortions, only about a fifth of physicians referred patients to a specific abortion provider (Holt et al., 2017). Among ob-gyns who do not perform abortions, one study found that only around half refer patients to specific facilities or clinics for abortions (Desai et al., 2018).

Lack of knowledge about where and whom to refer patients for abortion care has been consistently identified as a barrier to physician referrals (Holt et al., 2017; Homaifar et al., 2017; Zurek et al., 2015). However, researchers have not fully examined knowledge gaps about where and to whom to refer patients to for abortions, especially in the context of physicians who are willing to participate in abortion care. While a rich body of literature has focused on “conscientious provision” of abortion, where providers participate in abortion care despite personal objections (Czarnecki et al., 2019; Harris et al., 2011; McLemore et al., 2015), we lack research on the providers who are willing to participate in abortion care but lack the knowledge to do so.

### 1.4 Current Study

This study will advance our understanding of physicians who are willing to participate in abortion care (in this case, those who are willing to consult on abortion cases) but lack the knowledge to do so by examining characteristics associated with not knowing whom to refer a patient to for abortion care. To examine this question, we use a unique study of the physician faculty at the University of Wisconsin School of Medicine and Public Health. Although prior studies on abortion referrals have largely focused on primary care physicians and ob-gyns as these specialties engage in reproductive healthcare most frequently, it is advantageous to learn about referral knowledge throughout medical specialties as patients seeking an abortion may be engaged in different healthcare settings at the time of a pregnancy diagnosis.

Furthermore, the survey’s focus on physicians working in Wisconsin makes it an ideal case to learn about knowledge barriers to abortion referrals in a state that is considered hostile toward abortion access (Nash, 2019). Wisconsin is one of twenty-six states with laws in place that would restrict or criminalize abortion if *Roe v Wade’s* federal protections are removed (Nash, 2019). Wisconsin mandates medically unnecessary counseling, waiting periods, and ultrasounds prior to abortion; limits abortion coverage in Medicaid and health insurance policies offered to public employees; prohibits medication abortion via telemedicine; and restricts abortion based on a patient’s age, stage of gestation, and type of medical facility (Guttmacher Institute, 2021a). These factors and others contributed to the closure of 40% of the state’s abortion facilities between 2009 and 2017, which coincided with significantly higher birthrates in counties experiencing the greatest distance increases to abortion healthcare (Venator & Fletcher, 2021). Anti-abortion legislators in Wisconsin have also attempted to restrict abortion training opportunities at the state’s only public medical school, which would cause UW’s residency program to fall out of compliance with the ACGME national standards (Associated Press, 2017; Paris, 2021; White, 2017). Given this restrictive environment, Wisconsin creates a fitting case study in healthcare providers’ knowledge of where and how to refer patients to abortion care.

## 2. METHODS

### 2.1 Data

To assess the factors associated with physicians’ ability to refer patients for abortion care, we used a cross-sectional survey of physician faculty at the University of Wisconsin School of Medicine and Public Health (Schmuhl et al., 2021). This web-based survey, administered in 2019, gauged physicians’ knowledge, attitudes, and referral practices regarding abortion care and policies. Additional information about the survey design has been described in detail elsewhere (Higgins et al., 2021; Schmuhl et al., 2021). While multiple factors inspired the survey, two especially pertinent ones here are as follows. First, the research team wanted to address a comparative lack of abortion research with physicians across medical specialties (Schmuhl et al., 2021). Second, investigators identified the need for greater understanding of physicians’ abortion-related attitudes, knowledge, and practices in state-based contexts in which abortion access is already significantly restricted, and where it would become illegal with the overturning of *Roe v. Wade* (Higgins et al., 2021).

In conjunction with experts at the University of Wisconsin Survey Center, researchers fielded the survey to all practicing physician faculty members (*N* = 1,357) at the Wisconsin School of Medicine and Public Health—the largest and only state-supported medical school in the state. The team worked to maximize participation by using best practices such as a motivating incentive structure (e.g., $5 bills enclosed in hard-copy study invitations), Web and mail mixed-mode methodology, and up to three reminder emails and a final article questionnaire distributed to initial nonresponders. The team collected responses from February to May 2019.

Of 1,357 distributed surveys, respondents completed and returned 913, for an adjusted response rate of 67%. Participants represented more than 20 medical specialties, and 94% said their patients include women of reproductive age.

Given our interest in examining systemic, rather than attitudinal barriers to abortion care, we decided to focus on survey respondents who indicated that they are willing to consult in the care of any patient seeking an abortion. There were 148 respondents who reported that they were “not at all” or “a little” willing to consult on abortion cases, and these respondents were excluded from the analytic sample. Our study sample therefore includes the 675 physicians who are “somewhat,” “very,” or “extremely” willing to consult in abortion care (out of an initial 859 respondents). By focusing on only those physicians willing to consult on abortion care, we can examine one aspect of physician’s practical ability to help while assuming that, at the baseline, the physician is willing to help patients with abortion care.

### 2.2 Dependent Variable

We created a binary variable representing abortion referral knowledge. Respondents were asked “If you had to refer a patient for abortion care, would you know whom to contact?” This variable was reverse coded so those who responded “yes” were coded as 0 and those who responded “no” were coded as 1.

### 2.3 Explanatory Variables

We focus on several potential factors associated with physician referral knowledge. First, we include a measure of self-reported relevant expertise. Respondents were asked to rate how relevant their medical expertise is to abortion on 5-point scale from “not at all” to “extremely.” In regression analysis, relevant expertise is treated as continuous.^2^ To measure medical experience, we include a measure of the years practicing medicine since residency. Respondents could select between one of six categories: (1) “less than one year,” (2) “one to five years,’ (3) “six to ten years,” (4) “eleven to fifteen years,” (5) “sixteen to twenty years,” or (6) “more than 20 years.” This measure is treated as continuous.^3^

Three additional measures capture respondent’s familiarity with abortion care. First, we include medical specialty. The survey asked respondents to select one of fourteen categories. Due to small sample sizes, we recoded the specialty variable into five categories: (1) surgery, (2) pediatrics, (3) family medicine, (4) obstetrics and gynecology (ob-gyn), or (5) other specialties. Second, we include a binary indicator for exposure to abortion care training. Respondents were asked “Did you get any exposure to abortion care during your medical education, such as in medical school, residency or fellowship, even if you did not participate?” Respondents who answered “yes” were coded as 1, and respondents who answered “no” were coded as 0. Third, we introduce an indictor whether the respondent is aware that Wisconsin state policy has made abortion services illegal at University of Wisconsin Hospital and Clinics, which we refer to as the UW abortion restriction. Respondents aware of this policy were coded as 1.

We also included a measure of the respondent’s abortion attitudes. An abortion attitudes scale was created by averaging 9 items: (1) How necessary is the care that abortion providers provide for women, (2) Abortion providers make a positive contribution to society, (3) If my child became a physicians, I would be proud if they offered abortion services, (4) Abortion providers are heroes, (5) How happy to help would you be if an abortion provider called you for consultation about… a mutual patient seeking abortion care, (6) How much do you trust the motivations of abortion providers, (7) Abortion providers should be ashamed of their work, (8) How skilled are abortion providers who work in free-standing clinics such as Planned Parenthood, and (9) Overall, would you say that complications from abortions receive more or less scrutiny than complications from other medical care. Items 2, 3 and 4 were reverse coded so that higher values indicate positive attitudes toward abortion. The scale ranged from 1 to 5, and the Cronbach’s α was 0.92.

Lastly, we include two measures of the sociodemographic characteristics of respondents. The survey asked respondents to report their gender identity as (1) male, (2) female, or (3) other gender. Because there was only one respondent who identified as having an “other” gender identity, this individual was dropped from analyses. Respondents were also asked about their racial identity and were asked for their racial and ethnic identification. Due to small sample sizes, we recategorized race into a “person of color” category, where respondents who identify as white only were coded as 0, while respondents who reported being Hispanic/Latinx, African American or Black, Asian, Native American or Alaskan native, Arab or Middle Eastern, Pacific Islander or another race were coded as 1.

### 2.4 Statistical Analyses

All analyses were conducted in Stata 16.1. Respondents missing data on at least one variable were excluded through listwise deletion (N=16). The analyses consisted of two parts: descriptive analyses and logistic regression. First, we calculate descriptive statistics to both characterize the sample and develop a descriptive understanding of the characteristics associated with knowing whom to refer patients to for abortion care. Second, we estimate a logistic regression model to analyze patterns in which physicians are able to refer patients for abortion care.

## 3. RESULTS

### 3.1 Descriptive Results

Just over half (53%) of all physicians willing to refer patients did not know whom to refer them for abortion care (see Table 1). Male physicians, who made up 52% of our sample, were less likely to know whom to refer patients to for an abortion compared to female physicians (61.8% vs. 42.4%; p<0.001). About 86% of the physicians in our sample identified as white, a proportion higher than the national average of around 65% (Association of American Medical Colleges, 2019). A higher proportion of physicians of color reported not knowing whom to refer patients to for abortion care compared to physicians who identified as white (64.6% vs. 50.5%; p=0.01).

**Table 1:** Descriptive Results, University of Wisconsin SMPH Faculty Attitudes About Abortion, 2019^a^ (N=674)

| Categorical Variables |  | N | % of Sample | % Low Referral Knowledge |
| --- | --- | --- | --- | --- |
| Abortion referral knowledge |  |  |  |  |
|  | Yes | 320 | 47.5% | — |
|  | No | 354 | 52.5% | — |
| Gender |  |  |  |  |
|  | Woman | 323 | 47.9% | 42.4% |
|  | Male | 351 | 52.1% | 61.8% |
| Race |  |  |  |  |
|  | White | 578 | 85.8% | 50.5% |
|  | Person of color | 96 | 14.2% | 64.6% |
| Abortion care exposure during medical education |  |  |  |  |
|  | Yes | 376 | 55.8% | 43.4% |
|  | No | 298 | 44.2% | 64.1% |
| Aware of UW abortion restriction |  |  |  |  |
|  | Yes | 179 | 26.6% | 20.1% |
|  | No | 495 | 73.4% | 64.2% |
| Specialty |  |  |  |  |
|  | Internal medicine | 181 | 26.9% | 60.8% |
|  | Surgery | 68 | 10.1% | 72.1% |
|  | Pediatrics | 71 | 10.5% | 33.8% |
|  | Family medicine | 99 | 14.7% | 7.1% |
|  | Obstetrics & gynecology | 31 | 4.6% | 3.2% |
|  | Other | 224 | 33.2% | 72.8% |
| Continuous Variables |  | Mean | Standard Error | Mean for Unable to Refer |
| Relevant expertise |  | 2.5 | 0.05 | 2.0 |
| Abortion attitudes |  | 4.4 | 0.02 | 4.3 |
| Years since residency |  | 4.0 | 0.06 | 3.8 |

<sup>a</sup>This table includes only those respondents who that they are “somewhat,” “very” or “extremely” willing to consult in the care of a patient seeking an abortion.

We observed notable differences in referral knowledge by factors associated with medical training and experience. Just over 55% of physicians in this sample were exposed to abortion care during their medical education, and those who were exposed were more likely to know whom to refer patients to for abortion care (χ^2^=28.68; p<0.001). Even still, around 45% of physicians in our sample with exposure to abortion during their medical training did not know to whom to refer patients. Not knowing whom to refer patients for abortion care was particularly prevalent among physicians in our sample who specialized in internal medicine or surgery (60.8% and 72.1%, respectively). One third (34%) of pediatricians did not know who to contact for an abortion referral, but very few family medicine (7%) and ob-gyn (3%) physicians reported not knowing whom to refer patients to for abortion care. Among physicians who fell into the “other” specialty, 73 percent did not know whom to make an abortion referral.

Physicians who reported higher relevant expertise were more likely to know whom to refer patients to for abortion care (μ_relative expertise_=2.0 for physicians who did not know whom to refer patients to; μ_relative expertise_=3.1 for physicians who knew whom to refer patients to; t = 12.94 p<0.001). Physicians who had been practicing medicine for fewer years since the residency were less prepared to make abortion referrals than those with more experience (t = 3.16; p=0.01). Furthermore, almost three-quarters of respondents (73.1%) were unaware that abortion services are illegal at UW facilities, and those respondents who were unaware of this restriction more often did not know whom to refer patients for an abortion (64.2% vs. 20.1%).

We limited our sample to physicians who were willing to consult an abortion case. The mean abortion attitude score was equal to 4.4 out of a maximum score of 5. While abortion attitudes were significantly lower between physicians who did not know whom to refer patients to (μ=4.3) compared to physicians with high referral knowledge (μ=4.5; t = 5.73; p<0.001), the magnitude of the difference between these two scores was relatively small (Δ=0.2).

### 3.2 Regression Results

Regression results indicate clear associations between abortion care experience, abortion attitudes and ability to refer patients for abortion care (see Table 2). As a reminder, these analyses focus on physicians who reported willingness to consult on abortion cases. We observed no significant differences between physicians who identified as male versus female. However, white physicians were significantly more likely to be able to refer patients for abortion care compared to physicians of color (p=0.01).

**Table 2:** Logistic Regression Results Predicting Referral Knowledge.

|  | Odds Ratio<br>(S.E.) |
| --- | --- |
| Male | 1.40<br>(0.29) |
| Race (person of color) | 2.44**<br>(0.81) |
| Relevant expertise | 0.59***<br>(0.06) |
| Abortion attitudes | 0.56***<br>(0.11) |
| Years since residency | 0.86*<br>(0.06) |
| Abortion care exposure during medical education | 0.67*<br>(0.13) |
| Aware of UW restriction | 0.22***<br>(0.06) |
| *Specialty (vs. Internal medicine)* |  |
| Surgery | 1.44<br>(0.50) |
| Pediatrics | 0.48*<br>(0.16) |
| Family medicine | 0.12***<br>(0.06) |
| Obstetrics & gynecology | 0.09*<br>(0.10) |
| Other | 2.00***<br>(0.48) |
| *N* | 674 |

Exponentiated coefficients; Standard errors in parentheses. \* $p < 0.05$ , \*\* $p < 0.01$ , \*\*\* $p < 0.001$

Physicians with greater relative expertise were better equipped to refer patients for abortion care. For each one-point increase in reported relative expertise, the odds of not knowing whom to refer patients decreased by a factor of 0.59 (p<0.001). This finding is further supported by the association between specialization and referral knowledge. Figure 1 displays the predicted probabilities, the probability of not knowing whom to refer patients to for an abortion while all other values are held at their mean, by medical specialty. The specialties most likely to be unable to refer patients were those specializing in internal medicine, surgery or in one of the “other” medical specialties. Compared to physicians specializing in internal medicine, pediatricians were about half as likely to not know whom to refer patients to (OR=0.48; p=0.03). As expected, based on which specialties are likely to include abortion care training in residency (Greenberg & Nothnagle, 2018; Steinauer et al., 1997, 2018), the specialties most equipped to refer patients for abortion care were those who specializing in family medicine or ob-gyn.

**Figure 1:**
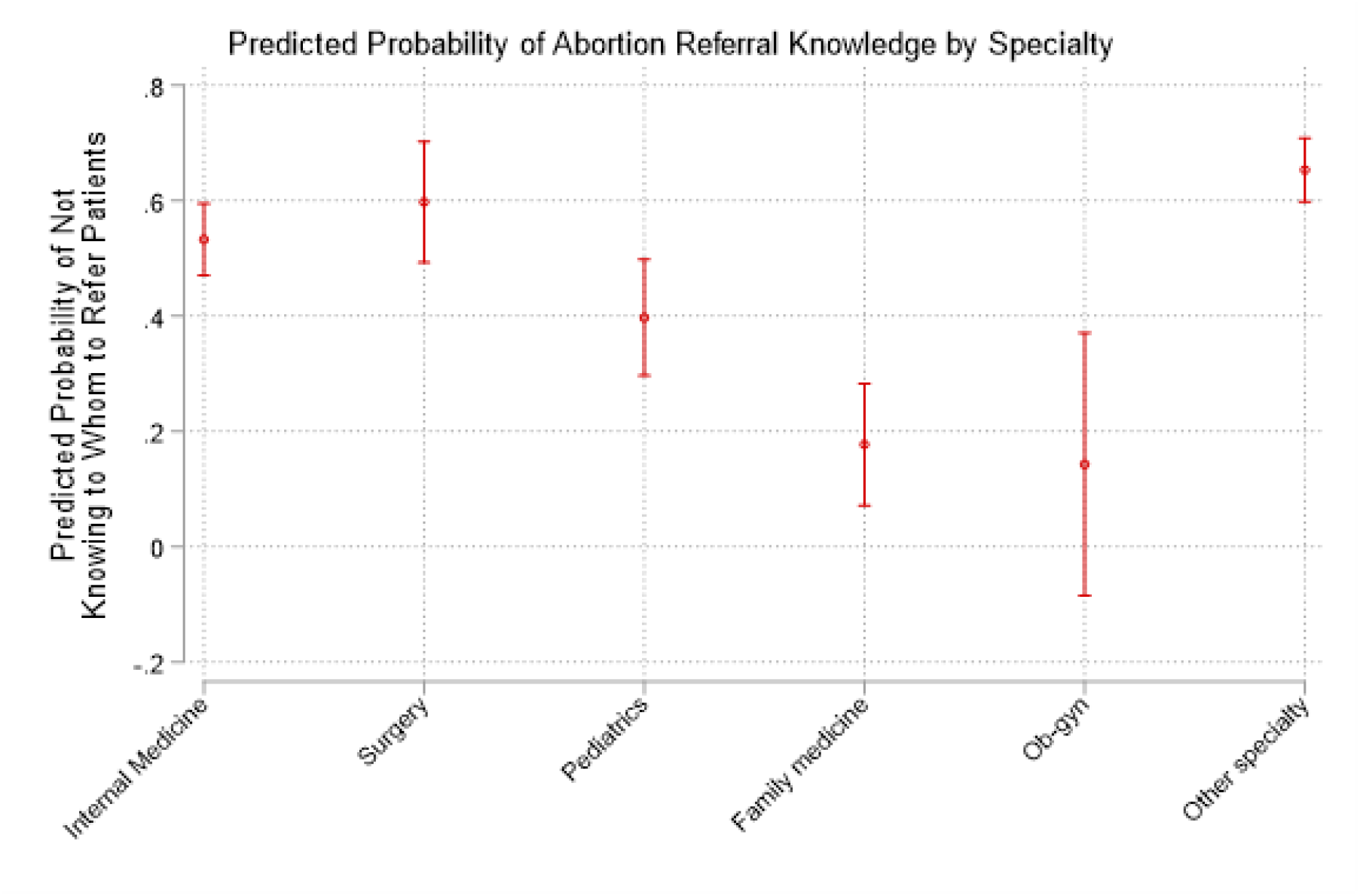
Predicted probability of abortion referral knowledge by specialty.

Physicians who were more supportive of abortion were less likely be unable to refer patients for abortion care. For each one-point increase in abortion attitudes, the odds not knowing whom to refer patients to decreased by a factor of 0.56 (p=0.002). However, because the physicians in this sample were, on average, very supportive of abortion, abortion attitudes in the sample likely does not substantively differentiate these physicians’ ability to refer patients for abortion care.

Aspects of medical training were also associated with ability to refer patients in important ways. Physicians who were exposed to abortion care at any point in their medical education were more likely to know whom to refer patients to for abortion care compared to those who were not exposed (OR=0.67; p=0.04). Overall experience as a physician was also meaningful as physicians who had been practicing medicine for more years after residency were also significantly less likely to not know whom to refer patients to for abortion care (OR=0.86; p=0.02). Furthermore, physicians who were aware of the state restriction on abortion care in UW facilities, which potentially indicates exposure to or experience with such care, were more likely to know whom to refer patients (OR=0.22; p<0.001).

## 4. DISCUSSION

Compelling previous research has examined “conscientious provision” of abortion, in which physicians may object to abortion personally but still participate in abortion care (Czarnecki et al. 2019; Harris et al. 2011; McLemore, Kools, and Levi 2015). However, scholars have yet to document whether physicians who are willing to participate in abortion care have the knowledge of how to refer patients for abortion services. Our findings highlight a sizable hole in the reproductive health referral system: half of the physicians in our sample who were willing to consult on abortion cases were unable to refer patients for an abortion. Although abortion is a primary healthcare procedure, even doctors who are willing to participate in abortion care lack the knowledge to effectively engage in a tangential role through referrals. Respondents’ exposure to abortion care, such as medical training in residency, as well as their years of experience as a physician, were both positively associated with physicians’ ability to refer. While this latter finding might seem unsurprising, at face level, it reflects the segregated nature of abortion within medicine in the United States and serves as an indicator of the collateral consequences of this segregation, especially among physicians with less experience. Regardless of specialty, every physician in this sample reported treating reproductive aged women.

This research builds on previous research on institutional constraints to ob-gyn physicians’ ability to provide abortion care (Freedman, 2010; Freedman et al., 2010) by examining a sample of physicians with a diverse range of specialties. Because past research on the provision of abortion care has largely focused on ob-gyn physicians, and, to a lesser extent, family medicine practitioners and nurses, the underlying assumption of this research presumes that the medical providers involved in abortion care have the requisite knowledge to participate in abortion care. While abortion training is not universal for any of these specialties, the healthcare providers are framed as being capable of providing care if not for institutional barriers or moral opposition.

In this work, we presume a level of willingness to participate in abortion care while examining knowledge (ability to refer patients for abortion care) as a potential barrier to abortion care. Abortion referrals are an important but understudied topic within research on barriers to abortion care. Although information about abortion services can be found online, referrals for abortion care can help address some of the barriers created by prolific misinformation online and scarce abortion facilities (Dodge et al., 2018; Kavanaugh et al., 2019). Legal restrictions continue to shorten the time frame during which abortions are permitted by law (e.g., the 2021 Texas law that bans abortion after six weeks of pregnancy). Consequently, individuals seeking an abortion must quickly mobilize resources and accurate, detailed information from healthcare providers can facilitate access (Kavanaugh et al., 2019). However, our findings indicate a widespread dearth of knowledge, indicating that this potential resource for individuals seeking an abortion is often unavailable.

Physician referrals are a common and important function of the United States’ medical system, as patients are regularly referred to specialists to address different healthcare needs (Barnett et al., 2012). Even if the need for an abortion referral is relatively unusual within a physician’s practice (Jones et al., 2019), all physicians should be equipped to refer patients for this common medical procedure. Interventions to address this knowledge gap have the potential to be an effective way to reduce one of the many barriers that people face when seeking an abortion. Incorporating training on how to refer patients to abortion care within medical residency programs or continuing medical education credits might mitigate the effect of inexperience on ability to refer. Furthermore, medical institutions as well as local medical associations could regularly provide information on how to refer patients for abortions so that physicians have access to accurate and timely referral information.

Our decision to focus on only those physicians willing to engage in abortion care through consulting in the care of a patient seeking an abortion, we sought to minimize the effect of personal beliefs about abortion and focus on the institutional forces shaping physicians’ ability to refer patients for care. Nevertheless, about a quarter (26%) of physicians who were unwilling to engage in abortion care knew whom to refer patients to for abortion care, indicating that referral knowledge is not wholly dependent on personal stance on abortion. This finding reflects previous research on the negotiated decisions that healthcare providers make regarding abortion (Czarnecki et al., 2019; Freedman, 2010; Harris et al., 2011; McLemore et al., 2015). However, the vast majority of physicians who refuse to consult on abortion cases were unable to refer patients for care, indicating that the absence of referral knowledge is potentially heightened by abortion attitudes.

### 4.2 Limitations and Strengths

While our unique dataset of physician faculty enabled us to examine abortion referral knowledge across specialty, our findings must be interpreted within several parameters. First, our sample size (N=647) is relatively small. To accommodate the sample size, we combined categories within the variables measuring physician race and medical specialty. In particular, a larger sample of physicians would enable an examination of more nuanced differences of referral knowledge between specialties. In addition, a larger sample of ob-gyn practitioners in the sample would also allow a direct comparison between other specialties and ob-gyn physicians. Second, our focus on physician faculty means that we cannot directly extrapolate our findings to all physicians and future studies should examine whether our findings generalize to physicians in private practice and other non-academic medical settings. Even after accounting for these limitations, this study’s unique collection of abortion attitudes and knowledge across specialties for physicians working in a state designated as hostile to abortion provides a platform to enhance our knowledge of abortion within the larger American medical system (Guttmacher Institute, 2021a).

### 4.3 Conclusion

People seeking abortions in the United States often face a multitude of barriers toward accessing care. We examined one institutional barrier to abortion access: physician knowledge of whom to refer patients to for abortion care among a sample of faculty physicians willing to engage in abortion care. We found that knowledge of whom to refer patients to for an abortion was relatively low and associated with exposure to abortion care during training, medical specialty, and experience. These findings illustrate the marginalized position that abortion holds within the American medical system, but they also point to potential low-burden interventions to decrease barriers to abortion care. Interventions to educate physicians of all specialties on local abortion access to facilitate referrals has the potential to facilitate easier access to abortion, even in the current climate of increasing abortion restrictions. Smaller scale interventions to increase physicians’ abortion referral knowledge should be paired with interventions at the level of medical school education and residency training to integrate training about abortion to develop a more robust referral system across specialties. Interventions to increase physicians’ knowledge about abortion care have the potential to create a robust abortion referral system, regardless of specialty, which would improve patient’s access to abortion care (Zurek et al., 2015).

## Data Availability

All data produced in the present study are available upon reasonable request to the authors.

## Acknowledgements

The authors would like to thank Lisa Harris, Lisa Martin, and Meghan Seewald at the University of Michigan for sharing their survey instrument and consulting on the content of our survey instrument. We are also grateful to the University of Wisconsin Survey Center for their invaluable methodological expertise, multiple reviews of the survey instrument, and data collection efforts.

## Appendix A

**Table A:**
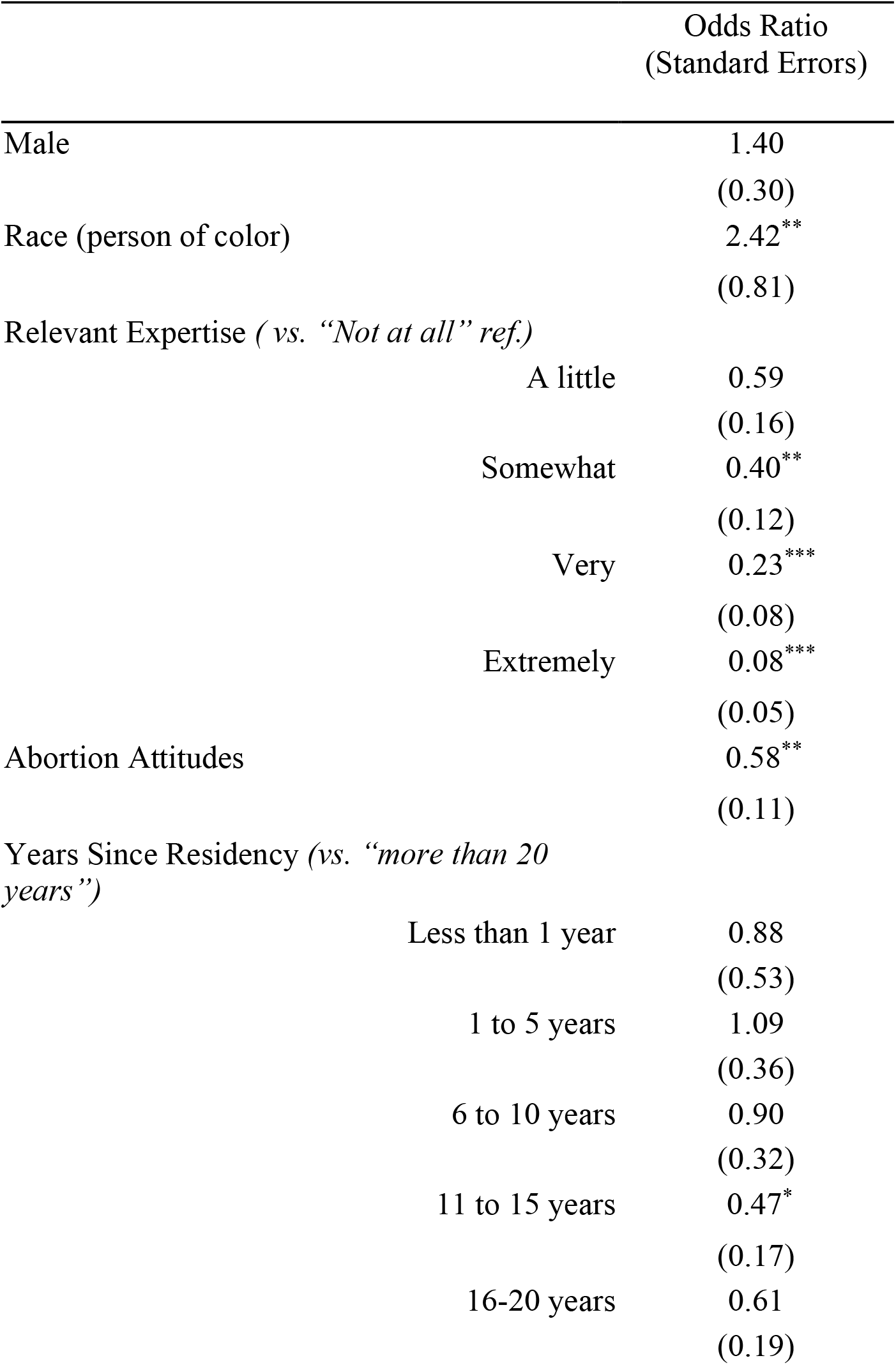
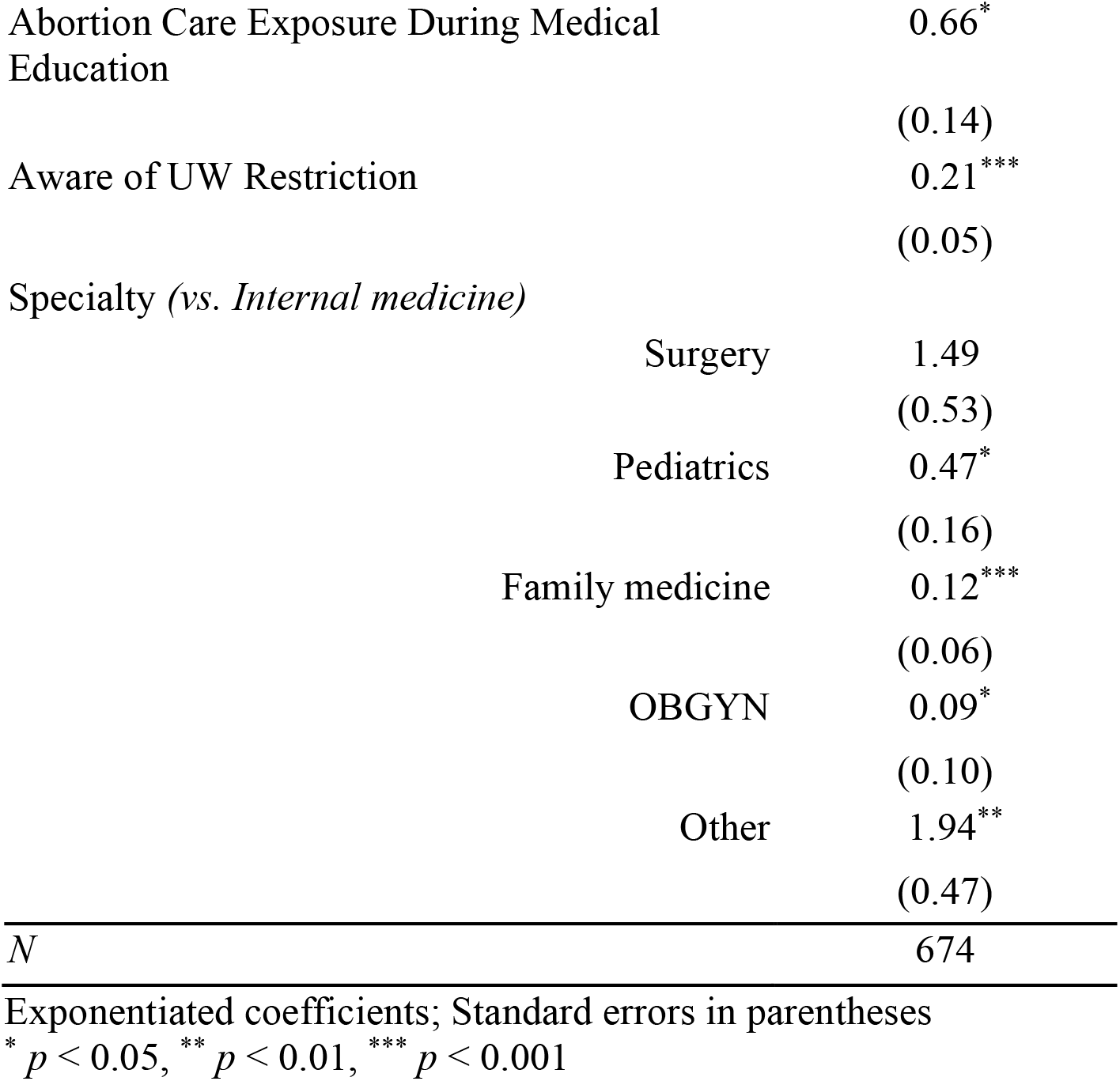
Sensitivity analyses with continuous specifications for relevant expertise and years since residency. Logistic regression predicting whether an individual who is willing to consult on abortion case is able to refer patients to abortion care (0 = willing to consult and able to refer, 1 = willing to consult but unable to refer).

## Footnotes

1 We used the term “women” in our questions because the overwhelming majority of abortion patients identify as women, but we note that trans men and gender-nonconforming individuals also need and seek abortions.

2 Sensitivity analyses using a categorical specification for relevant expertise produces results that are largely consistent with the continuous specification (see Appendix A).

3 Sensitivity analyses using a categorical specification produces non-significant results. However, we chose to use the continuous specification to indicate the effect of cumulative experience over time.

